# Data Quality in health research: a systematic literature review

**DOI:** 10.1101/2022.05.31.22275804

**Authors:** Filipe Andrade Bernardi, Domingos Alves, Nathalia Yukie Crepaldi, Diego Bettiol Yamada, Vinícius Costa Lima, Rui Pedro Charters Lopes Rijo

## Abstract

Decision-making and strategies to improve service delivery need to be supported by reliable health data to generate consistent evidence on health status, so the data quality management process must ensure the reliability of the data collected. Thus, through an integrative literature review, the main objective of this work is to identify and evaluate digital health technology interventions designed to support the conduct of health research based on data quality. After analyzing and extracting the results of interest, 33 articles were included in the review. This transdisciplinarity may be reaching the threshold of significant growth and thus forcing the need for a metamorphosis of the area from focusing on the measurement and evaluation of data quality, today focused on content, to a direction focused on use and context

In general, the main barriers reported in relation to the theme of research in the area of health data quality cite circumstances regarding a) use, b) systems and c) health services.. The resources presented can help guide medical decisions that do not only involve medical professionals, and indirectly contribute to avoiding decisions based on low-quality information that can put patients’ lives at risk

## Introduction

In health, the relevance and performance of digital media are evidenced by the efforts of governments around the world to develop infrastructure and technology aiming to expand their ability to take advantage of generated data. It is important to emphasize that technology, by itself, is not capable of transforming data into information, and human participation is essential for the production of knowledge from a set of data. Through research that optimizes health interventions and contributes to the alignment of more effective policies, knowledge combines concrete experiences, values, contexts, and insights, which may enable a framework for evaluation and decision making [1].

The low quality, availability, and integration of health data can be highlighted among the main factors that negatively influence research and health decision-making. In addition, it is worth noting the existence of a large number of isolated databases accessible only in a particular context. Such factors cause data quality problems and, consequently, information loss. Despite the intense volume, information remains decentralized without being able to help the decision-making process [2], making its coordination and evaluation challenges.

For example, the use of poor-quality data in the development of Artificial Intelligence models can lead to decision-making processes with erroneous conclusions. Artificial Intelligence systems, which are increasingly being used to aid decision-making, have used labeled sets of big data to build their models. Data is often collected and labeled by low-paid algorithms, and, increasingly, research demonstrates the problems of this method. Algorithms can present biases in judgments about a person’s profession, nationality, or character, and basic errors hidden in the data used to train and test their models. Consequently, prediction can be disguised, making it difficult to distinguish between right and wrong models [3].

The heterogeneity of data in this area is intrinsically connected to the type of information generated by health services and research, which, in themselves, are considered diverse and complex. The highly heterogeneous and sometimes ambiguous nature of medical language and its constant evolution, the enormous amount of data constantly generated by process automation and the emergence of new technologies, and the need to process and analyze data for decision-making constitute the foundation for the inevitable computerization of health systems and research and to promote the production and management of knowledge [4].

There are different concepts for “data quality” [5]. According to the World Health Organization, quality data portrays what was determined by its official source and must encompass the following characteristics: accuracy and validity, reliability, completeness, readability, timeliness and punctuality, accessibility, meaning or usefulness, confidentiality, and security [6]. Data quality can be affected at different stages, such as the collection process, coding, and non-standardization of terms. It can be interfered with by technical, organizational, behavioral, and environmental aspects [7].

Even when data exist, some aspects make their use unfeasible by researchers, managers, and health professionals, such as the non-computerization of processes, heterogeneity, duplicity, and errors in collecting and processing data in health information systems [8]. Decision-making and strategies to improve service delivery need to be supported by reliable health data to generate consistent evidence on health status, so the data quality management process must ensure the reliability of the data collected [9].

Some health institutions have action protocols that require their departments to adopt quality improvement and resource savings initiatives. Consequently, various methodologies to improve the quality of services have been applied in the health field. Likewise, research in scientific communities about new strategies is constantly evolving to improve research quality through better reproducibility and empowerment of researchers and patient groups with tools for secure data sharing and privacy compliance [10].

In order to use best practices, institutions present research related to review methods due to the significant time savings and high reliability of their studies. Thus, through an systematic literature review, the main objective of this work is to identify and evaluate digital health technology interventions designed to support the conduct of health research based on data quality.

## Methods

To define the research question, the Population, Concept, and Context (PCC) strategy was applied. It is a strategy that guides the question of the study and its elaboration, helping in the process of bibliographic search for evidence. The adequate definition of the research question evidences the information necessary to answer it and avoids the error of unnecessary searches [11].

Population refers to the population or problem to be investigated in the study. Content refers to all elements detailed and relevant to what would be considered in a formal systematic review, such as interventions and phenomena of interest and outcomes. Context is defined according to the objective and the review question. It can be determined by cultural factors such as geographic location, gender, or ethnicity [12]. For this study, the following were defined: P - *Digital Technology*, C - *Data Accuracy*, and C - *Health Research*.

In this sense, the following research questions were defined:

- What is the definition of health research data quality?
- What are the health research data quality techniques and tools?
- What are the indicators for data confidence level in health research?

### Health research

#### Studies designs

Numerous classifications characterize scientific research depending on its objective, type of approach, and nature. Regardless of the purpose of how surveys can be classified, levels of confidence in data quality must be ubiquitous at all stages of the survey. Detailed cost-effectiveness analysis may inform decisions to adopt technology methods and tools that support electronic data collection of such interventions as an alternative to traditional methods.

Health research systems have invested heavily in research and development to support sound decisions. In this sense, all types of studies were observed that presented results of recent opportunities to apply the value of digital technology to the quality of the information in the direct or indirect evaluation in the promotion of health research. Therefore, in a transversal way, we considered all types of studies that described how to deal with such aspects.

#### Types of participants

Various methods for setting priorities in health technology research and development have been proposed, and some have been used to identify priority areas for research. This includes surveys and measurements of epidemiological estimates, clinical research, and cost-effectiveness assessments of devices and drugs. The technical challenges and estimation of losses due to variations in clinical practice and deviations from protocols have been supported by recommendation manuals and good practice guidelines. However, each of these proposed methods has particular severe methodological problems.

First, all these approaches see research simply as a method of changing clinical practice. However, there are many ways to change clinical practice, and conducting research may not be the most effective or cost-effective way. The real value of research is to generate information about what clinical practice should be. The question of how to implement the survey results is a separate but related issue. Therefore, these methods implicitly assume no uncertainty surrounding the decision that the proposed research should inform.

#### Types of interventions

Technology-based interventions that affect and aggregate concepts, design, methods, processes, and outcomes promote data quality from all types of health research studies.

#### Types of evaluated results

Measures to demonstrate how results can address political, ethical, and legal issues, including the need to support and use technological mechanisms that bring added value regardless of the type and stage at which they are applied to research. We also look at how the results can be evaluated to address other questions, such as which subgroups of domains should be prioritized, which comparators and outcomes should be included, and which follow-up duration and moments would be most valuable for improving interventions on the reliability of health research data.

#### Eligibility criteria

Research carried out in English and Portuguese, with a quantitative and qualitative approach, primary studies, systematic reviews, meta-analyses and meta-synthesis, books, and guidelines, published from 2016 onwards were included. The year limitation is justified because knowledge is considered to have an adequate degree of up-to-dateness.

In addition to the methodological design, we adopted the inclusion criteria for any studies that describe the definition, techniques, or tools that have essential functions of synthesis, integration, and verification of existing data from different research sources to guarantee acceptable levels of data quality. In this way, we expect to monitor trends in health research, highlight areas for action on this topic, and, finally, identify gaps in health data arising from quality control applications.

While the primary objective of this review is to seek evidence of data quality from health research, we also independently included studies on health data quality and research data quality. The exclusion criteria were applied to studies with a lack of information (e.g., the article was not found); studies whose primary focus is not health and research; and articles not relevant to the objective of the present research; Also, the titles and the respective authors were checked to verify possible repetitions in the databases.

#### Databases and search strategies

A search was carried out in six electronic scientific databases in January 2022 at: PubMed, SCOPUS, Web of Science, IEEE Digital Library, CINAHL, and LILACS. For the search, descriptors and their synonyms were combined according to the Health Sciences Descriptors (DeCS) [13] and Medical Subject Headings (MeSH) [14]. The following descriptors and keywords were selected: Data Accuracy, Data Gathering, and Health Research, which were combined through the Boolean connectors AND and OR. The same search strategy was used in all databases. The Google Scholar database was used for manual searching and searching references of thesis and dissertations in the so-called gray literature.

A list of all studies found was created, and duplicates were removed. A manual search was performed to search for possible studies/reports that were not found in the databases. The references of each analyzed study were also reviewed for inclusion in the search. The search was carried out in April 2021, and based on the inclusion and exclusion criteria described, we arrived at the final number of articles included in the proposed integrative review. The search procedure in the databases and data platforms is described in Table 1, according to the combination of descriptors.

**Table 1 –.**
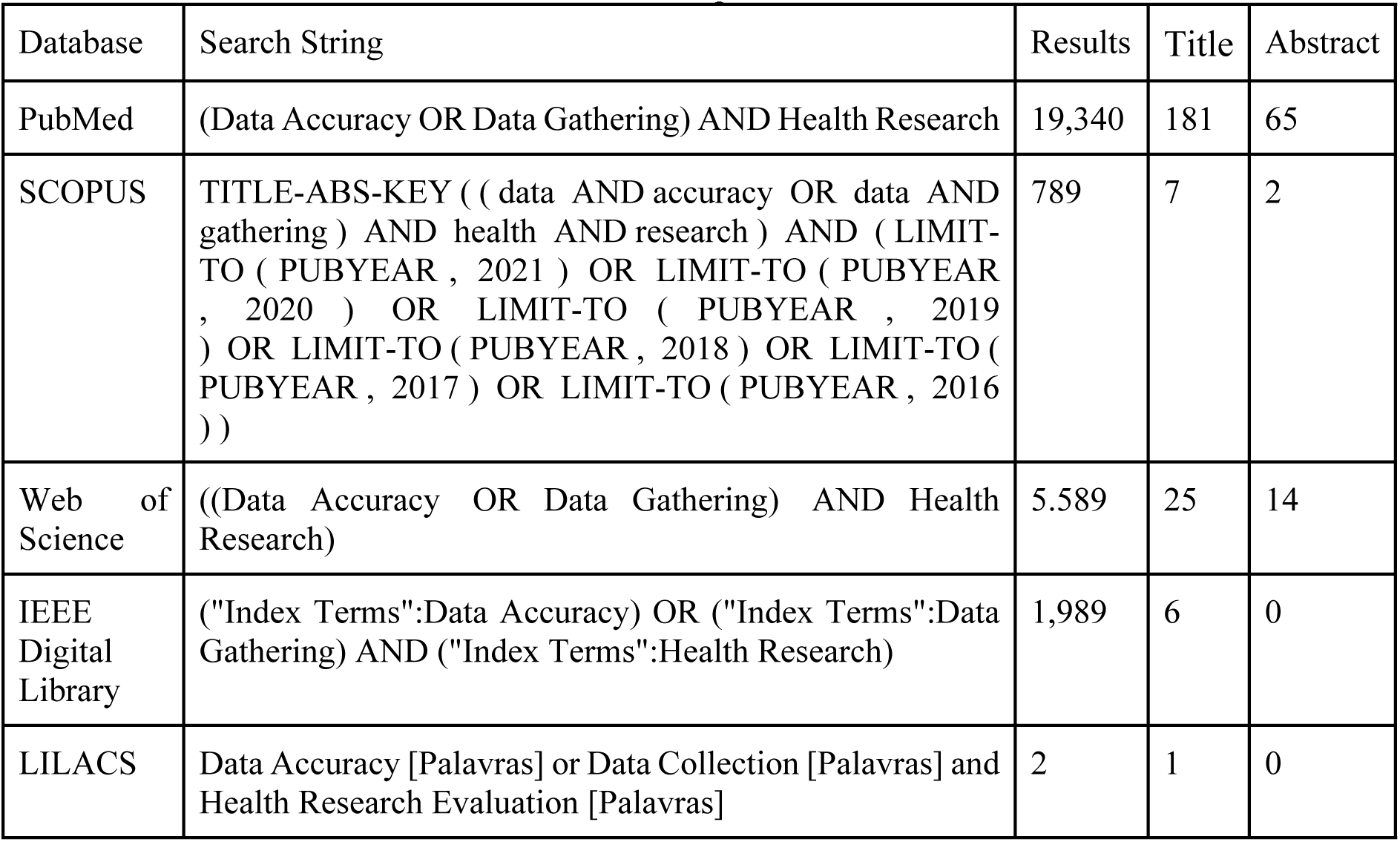
Search procedure on databases

#### Selection of studies

Data extraction involves first-order (participants’ citations) or second-order (researcher’s interpretation, statements, assumptions, and ideas) concepts in qualitative research. Second-order concepts were extracted to answer the questions of this research.

#### Data collection process

To identify the studies, a careful reading of the title, abstract, and keywords of all publications located by the search strategy was performed, and later, their suitability to the established inclusion and exclusion criteria was verified. The Mendeley reference manager was used to organize the articles [15]. Subsequently, the extracted findings were shared and discussed with the other team members.

Data synthesis aims to gather findings into themes/topics that represent, describe, and explain the phenomena under study. The extracted data were analyzed to allow a clear identification of themes arising from the data and facilitate the integration and development of the theory. Two reviewers performed data analysis and then shared it with other team members to ensure that the synthesis adequately reflects the original data.

#### Data extraction

We looked at data quality characteristics in the studies examined, the assessment methods used, and basic descriptive information, including the article and type of data under study. Before starting this analysis, we looked for pre-existing data quality and governance models specific to health research but could not find any.

Thus, two reviewers were responsible for extracting the following data from each article:

- Bibliographic information (title, publication date and journal, and authors);
- Study objectives;
- Methods (study design, data collection, and analysis);
- Results (researcher’s interpretation, statements, assumptions, and ideas).

#### Results presentation

The PRISMA (Preferred Reporting Items for Systematic Reviews and Meta-Analyses) checklist and flowchart were used for graphical visualization of the results of the search strategy in the databases. PRISMA follows a minimum set of items to improve reviews and meta-analyses [16]. Based on the PRISMA flowchart, a narrative synthesis was prepared in which we described the objectives and purposes of the selected and reviewed documents, the concepts adopted and the results related to the theme of this review.

## Evidence synthesis

### Studies Characteristics

In this review, 27,709 occurrences were returned from the search procedure, with 709 records from the SCOPUS database, 2 from LILACS, 1,989 from the IEEE digital library, 5,589 from the Web of Science, and 19,340 from Pubmed. Searches were also performed in the WHOLIS and CINAHL databases, but no results were found. Of these, 25,202 records were flagged as ineligible through the automation tools and filters available in the databases. Such items were mainly reports, editorial articles, letters or comments, book chapters, dissertations, and thesis, or because they did not specifically address the topic of interest and according to the use of descriptors. Furthermore, 204 records were duplicated between bases and were removed. We detailed and illustrate the process of inclusion and classification data from our findings in supplementary files.

After careful evaluation of the title and abstract (first screening step), 1221 search results were excluded. For inclusion of articles after reading the abstracts, 81 articles were listed for a full reading. After analyzing and extracting the results of interest, 33 articles were included in the review because they answered the search question. The entire process of selection, sorting, extraction and synthesis are described through the PRISMA flowchart [16] represented in Figure 1.

**Figure 1 -.**
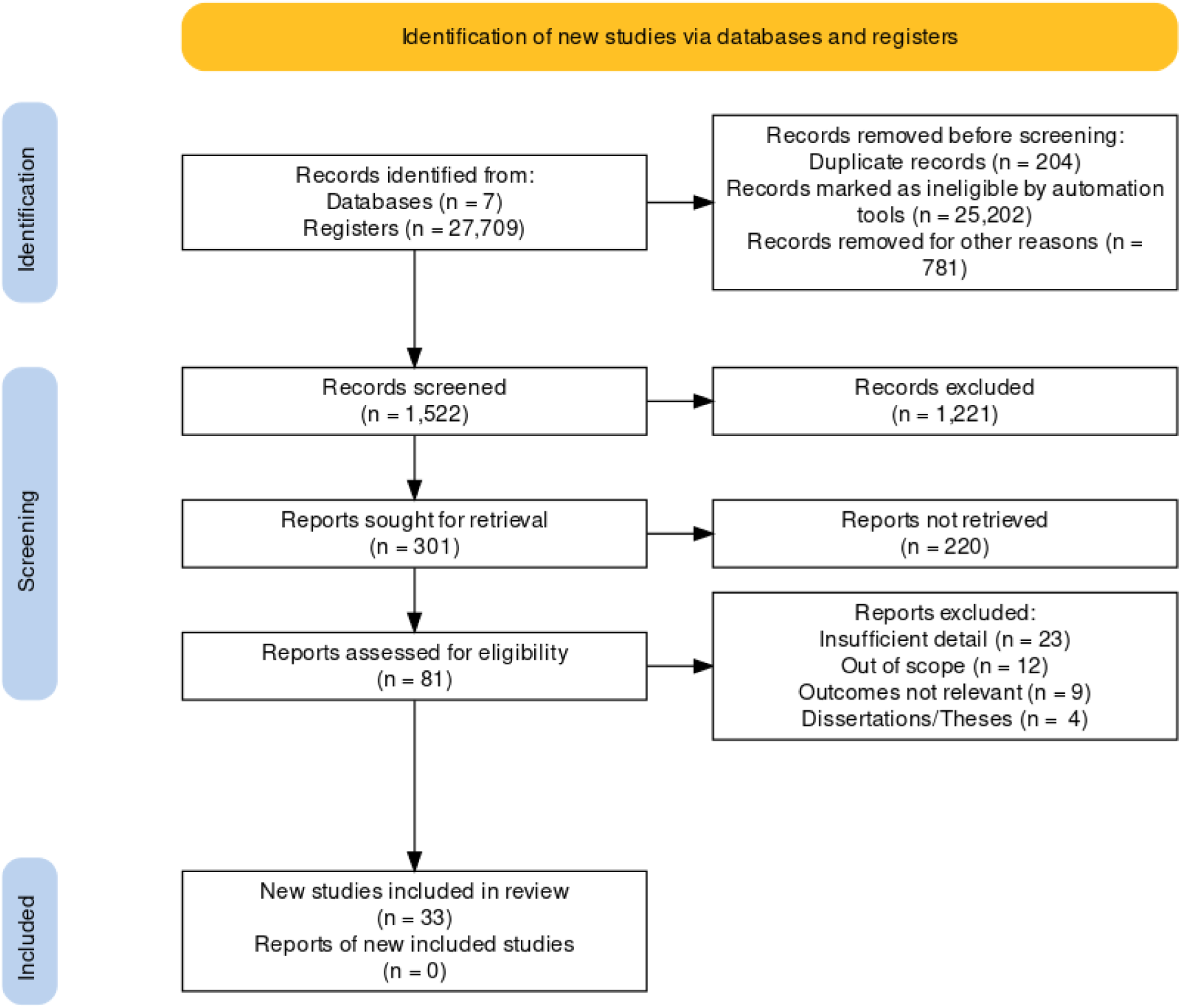
PRISMA flowchart with the results of studies selection Source: Prepared by the author.

The studies covered the years 2017 to 2021 and involved 22 countries. Of the 33 studies, 11 were concentrated in Europe, 10 were in North America (United States of America - USA and Canada). Another 4 were carried out in Oceania (Australia), 3 in Asia (China and Taiwan), and 2 in the Middle East (Iran, Saudi Arabia). In addition, 3 studies carried out their work collaboratively and/or in a network (USA and India; USA and countries on the African continent; US Consortium, UK, South Africa, Costa Rica, Canada, Sweden, Switzerland, and Bahrain).

In their entirety, the studies were carried out in developed countries, and most of the assessments were carried out based on the evidence available in the English language. The USA and Australia lead in studies involving the investigated topic, with 11 and 4 publications, respectively. No studies conducted or coordinated by emerging countries were reported. In addition to the low economic diversity of countries where the research was conducted, all articles were evaluated in a single language. The involvement and collaboration of emerging countries took place exclusively through partnerships and participation in consortia.

The metrics extracted from the studies comprised domains related to the methodology adopted by them, that is, which concepts supported the definition of data quality and their respective individual or combined categorizations, regarding the adjusted use for the purpose (8), of frameworks (6), ontologies (2) or good practice guides (15). The study design was also taken into account when defining the methodological proposal. In this scenario, there is a predominance of longitudinal studies (27), while cross-sectional studies (3) and/or studies that combined both methodological designs (2) were present in smaller numbers.

The environment, application time, and development of each study were considered. That is, if there was an evaluation of the evolution of the proposed solution from conception to its application. In a controlled environment in a research-only scenario, the planning and proof of concept development were reported in 7 studies, while the transition and validation in a real-world environment, where research and service are combined, were described in 8 opportunities. The use restricted to routine and occurred only in health services, due to some specific need in the area, were presented in 18 results. Most of these studies used their research repositories (24), while only 6 were based on pre-existing data models (4) or used public databases (2).

Many of the strategies and interventions used in these scenarios and data sources were performed with the objective of planning, managing and analyzing the impact that the implementation of at least one or more procedures has on data quality assurance. Most studies described well-established processes (22), 12 of which adopted models from the area of business intelligence, while another 7 used monitoring strategies, very common in the performance of clinical studies. In turn, 3 studies chose to use a process, originating from the commercial area, to search for best practices based on competitiveness (benchmarking) in order to obtain better evaluation results. On the other hand, 11 studies did not describe following a well-established process in the quality control of their studies,

The techniques and analyzes used in our findings obtained a great balance in relation to the type of method adopted, being 12 of them quantitative (12) and 11 qualitative, in addition to 10 studies that combined both. In addition, tools for defining a minimum set of data (7), auditing (13), error detection (3), and decision support (2) were also described. Two other studies used more than one tool to perform, respectively, the definition of the minimum set and auditing, and auditing and error detection. Finally, the other 6 did not report using any tools.

The main limitations of the studies recognized by the authors reflected methodological difficulties (21); techniques (15); social (6); organizational (2) and legal (1). Regarding the domains described in the studies, there is great variability and inconsistency between the terms presented (38 in total). Note that there is no consensus between critical and non-critical variables for data quality assessment. This also reflects that the definitions of concepts are varied and their relationships are not homogeneous across studies. Thus, the discrepancy between domains and evaluated concepts does not allow an evaluation of parity between metrics and is present during all phases of the studies found here.

Figure 2 depicts the great diversity of elements involved in the data quality process in health research, representing the planning (pre-collection), development (data acquisition and monitoring) and analysis (post-collection) stages. In our findings, each of the phases presented a set of strategies and tools implemented with a focus on providing resources that helped the interaction between the phases.

**Figure 2 -.**
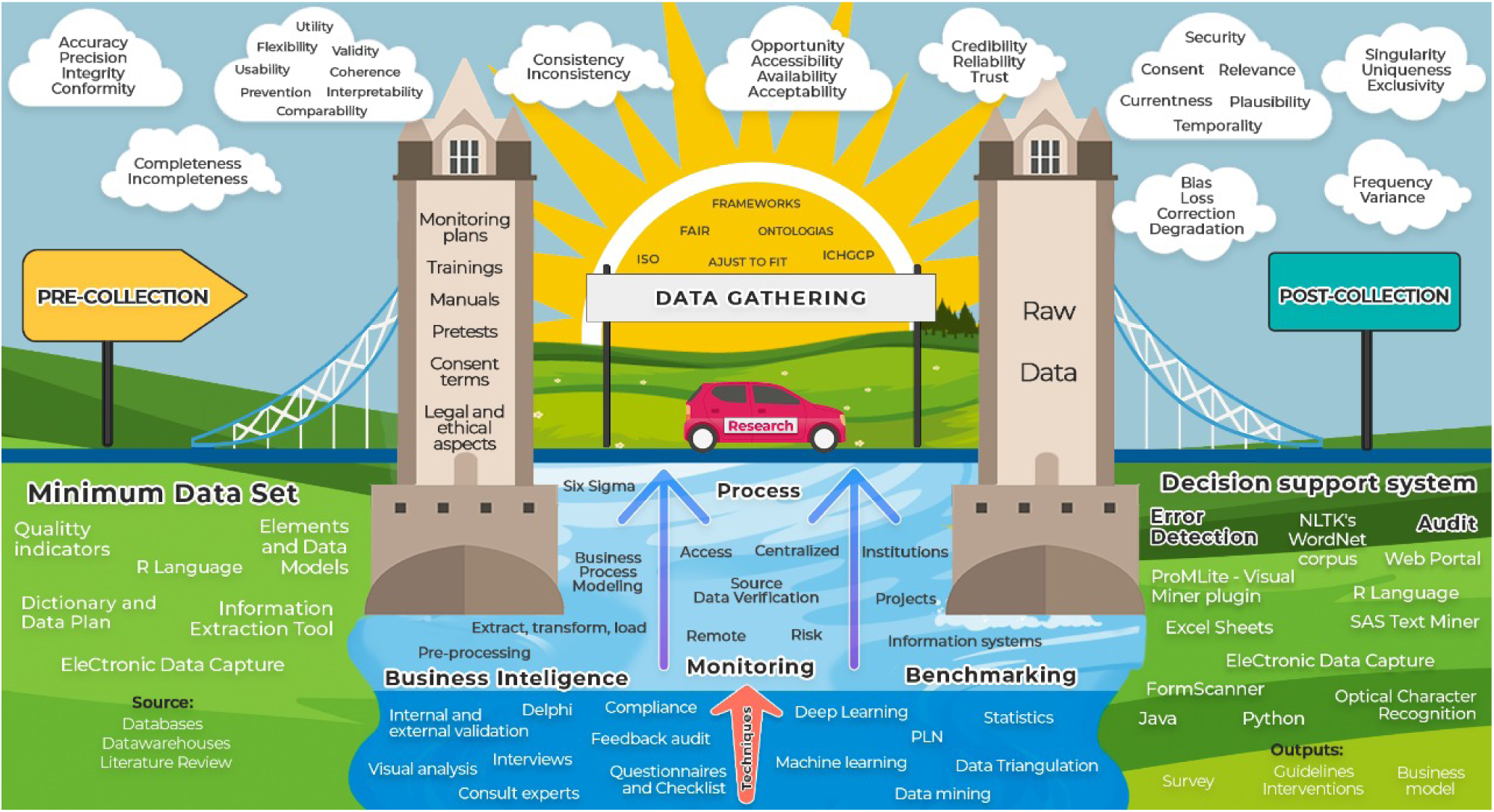
Elements involved in the research data quality process Source: Prepared by the author.

For the success of the research, the processes and techniques must be fluid and applied in a direction based on good guides and recommendations. Thus, the research must go through phases with well-established bases and tools, suitable for its purpose, making use of sources and instruments available through digital strategies and systems, as well as models, guides and feedback, and audit mechanisms.

In addition, every beginning of a new phase must be supported by well-defined pillars that encompass the exhaustive use of validations and pre-tests, plans for monitoring, management, and data analysis, precautions for ethical and legal issues, training of the team, and channels of effective communication. In terms of data quality, dimensions are interposed in all stages of research, being thus also a fundamental factor to being incorporated together with good practices and recommendations, giving light to health research, regardless of their methodological designs.

In the broadest sense, the process of incorporating data quality techniques and tools is analogous to going on a trip, that is, going from point A to point B. The starting point refers to good planning of issues such as the season of the year, the quantity and type of items that will be transported, the most appropriate means of transport, the budget available, as well as tips and guidance available in the different means of communication. Even if the path is already known, an important step that precedes the beginning of its execution is always the definition of the best route, and, for that, consulting maps and updated conditions are always recommended, since they can change throughout time.

On the other hand, the execution phase of a trip is not limited to reaching the final destination, during its journey, always being attentive to the signs and directions, without obviously failing to enjoy the landscape and all its opportunities, which together can provide greater entertainment. Finally, when we arrive at our destination, it must be borne in mind that to obtain the best results it is necessary to know the best guides and tourist attractions since a wrong choice or decision can provide us with a low-quality photograph, an unexpected experience. and as an effect, get an epilogue of bad memories.

### Methods

Among the studies that used the concept of purpose-adjusted use, terms such as “intrinsic quality”, “gold standard according to experts”, “ideal record”, “data fitness” and “data culture” were addressed. In general, the use of frameworks and ontologies was based on previously published studies and available in development libraries as modules for mapping adapted entities, proprietary and/or embedded systems, as well as data-based strategies for process improvement.

The main guides and guidelines adopted in data quality studies refer to the adoption of national protocols and policies, agreements signed between research networks and consortia, guides to good clinical practices (International Conference on Harmonization-Good Clinical Practice - ICHGCP; Food and Drug Administration - FDA; Health Insurance Portability and Accountability Act - HIPPA), and/or information governance principles, models and strategies (International Organization for Standardization - ISO; Joint Action Cross-Border Patient Registries Initiative - PARENT; Findability, Accessibility, Interoperability, and Reuse - FAIR).

The longitudinal studies are distributed in 22 retrospectives, where 5 were literature reviews and 17 were carried out in databases. All 5 prospective studies were carried out through surveys. Cross-sectional studies were presented in the form of clinical trials, business models and intervention studies.

### Participants

The diversity of health application areas were diverse 18 (Electronic health record (2); Cancer (2); Intensive care unit (ICU) (1); Rare diseases (RD) (2); General (4); Hospital (3); Neurodegenerative disease (1); Maternal health (1); Urology (1); Malaria (1); Emergency (1); Autoimmune disease (1); Cardiac (2); Emergency (1); Epidemiological (1); Outpatient (1); Immunization and Surveillance (1))). Meanwhile, the research areas are concentrated in fewer specialties 6 (Clinical research (5); Health informatics (2); Research networks (5); Social networks (1); Health learning system (1)); Clinical trials (1)). The health areas with the greatest applications encompassed applications that involved research fields in general (4) that took place in the hospital environment (3), with a greater number of participants (countries and/or institutions) in the RD area (2).

In the research scenario, the participation of collaborative research networks (5) and clinical action (5), that is, the one with the objective of evaluating the safety and efficacy of a procedure or medication, were predominant of the application areas, the application format focused on data sources that mainly reflected: sets of articles extracted from the literature (6), own repositories (11), based on institutional records (7), sets of clinical documents (1), the perception of experts (1) or data models (1), simulation models (1) and government databases (1).

### Interventions

For studies that define a strategy based on a business intelligence model, 5 followed the extraction, transform, load approach; 5 some form of preprocessing; 2 adopted the Six Sigma set of practices, while 1 adopted the Process Management Model (BPM). Data monitoring was carried out with approaches: risk-based (4); through data source verification (3); centrally (2); remotely, such as telephone contact (2); or through training (1). In addition, two studies applied the benchmarking strategy across systems (2), while another study used the same method across projects.

The vast majority of quantitative analyses adopted the combined use of strategies of the same nature. Data triangulation (3), for example, was always combined with statistical analyses, which in turn were used in other 3 studies in combination with data mining techniques (1), deep learning (1) and natural language processing (NLP) (1). In another 8 studies, statistics were used without any combination, thus making up the most used quantitative technique (14). The data mining technique was combined with NLP on one occasion. The use of data mining in isolation was reported on another occasion, as well as a machine learning model [17] and the application of the Latent Semantic Analysis (LSA) technique of NLP [18].

On the other hand, the qualitative analyzes were diverse (9 types) and combinations of the same nature were not used, with emphasis on consultation with specialists (6), and the use of structured instruments (4). The internal and external validation of the dataset (3) and the use of visual analysis (3) were also described in 6 studies. The use of methods such as interview, Delphi technique, feedback audit, use of grammatical rules, and compliance enforcement was also reported.

To perform such analyzes and processes, different computational resources were used. Among the programming languages, the R language was the only one used for planning and defining data sets (1), while for auditing databases and studies, in addition to the R language (3), one study reported the use of Python. For error detection, an occurrence of the R language and another of the Java language are described. For support and decision support, the use of clinical and administrative software was described, as well as a web portal, without any mention of the languages used.

Furthermore, the use of several electronic data capture platforms (EDC) have been described (REDCap; CommonCarecom; MalariaCare; AP-HP-CDR; CIS) for the purpose of planning and collecting the dataset (2) and auditing them (4). Still referring to the minimum data set strategy, the use of dictionary and data plan (3) and quality indicators (2) were described. Finally, the audit tools included the definition of a data monitoring plan, an electronic measurement (eMeasure), and the use of Excel spreadsheets.

### Results

Although 8 studies did not report any type of limitation in their development, 14 showed that data quality assessment suffers from at least two types of challenges. The technical limitations reported the absence and need to compare their methods (11); performance concerns in real-world scenarios (4); and the difference between the adopted infrastructure levels (2). Lack of effective methods of tracing IP addresses to security measures (1) and adequate visualization methods (1); the dependence on rules (1) and data types in a given context (1); and access and manipulation of data sources (1) were also reported as technical factors of attention.

Other aspects such as the disparity between the levels of knowledge of professionals (3) who participate in the research and assess the quality of data and the human inability to process and analyze large volumes of information (3) were mentioned. The absence of human (1) and material (1) resources to perform such activities was also reported. The specific legal limitation was due to differences in organizational policies that did not allow for the expansion of extensive analysis.

The main challenge that the studies reported refers to methodological approaches, especially regarding the impossibility of evaluating their solutions in more than one scope (13), with a more adequate sample size (10) and with cuts in different periods (3). The absence of a gold standard (1), the non-validation of developed models and the evaluation of methods in only one study design (1) are also described as possible barriers in the findings.

Finally, the distribution of dimensions evaluated in our findings showed great heterogeneity and is shown in Figure 3.

**Figure 3 -.**
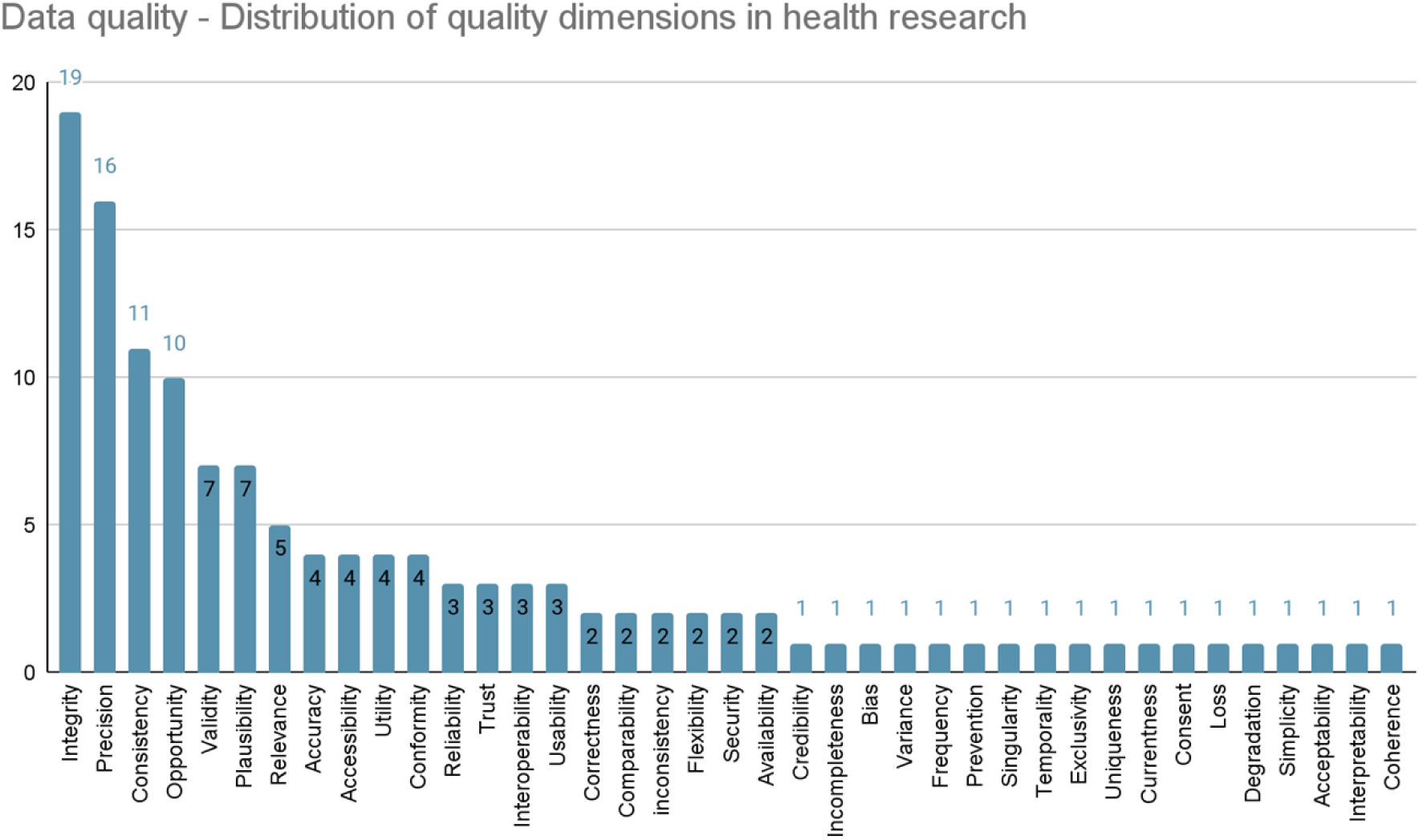
Distribution of quality dimensions in health research Source: Prepared by the author.

### Summary of findings

In general, the main barriers reported in relation to the theme of research in the area of health data quality cite circumstances regarding a) use, b) systems and c) health services. Such barriers are influenced by technical, organizational, behavioral, and environmental factors that cover large contexts of information systems, specific knowledge and multidisciplinary techniques [19]. The quality of each data element in the nine categories can be assessed by checking their adherence to institutional norms or by comparing and validating them with external sources [20]. Table 2 summarizes the main types of obstacles reported in the studies.

**Table 2 -.**
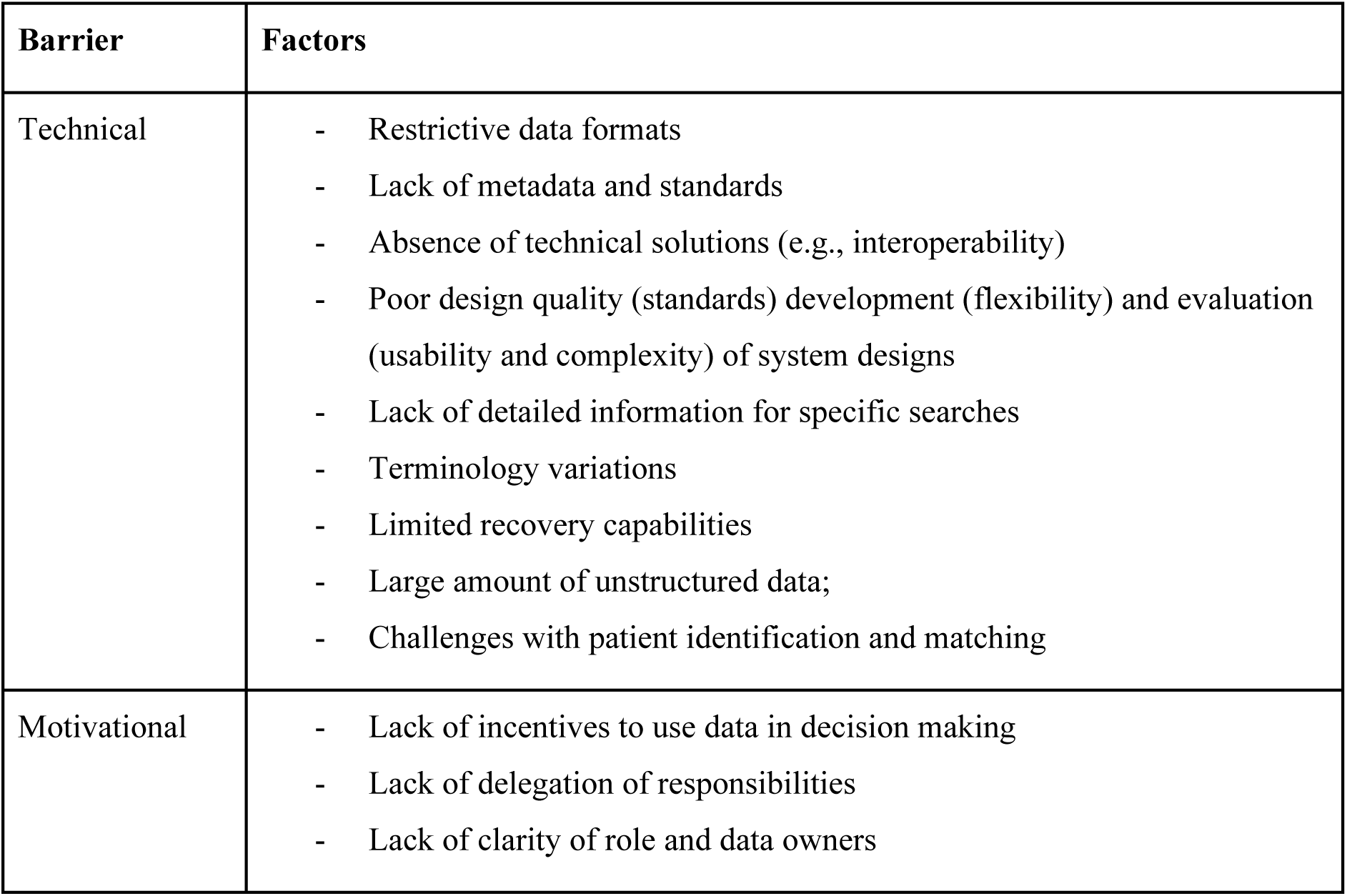
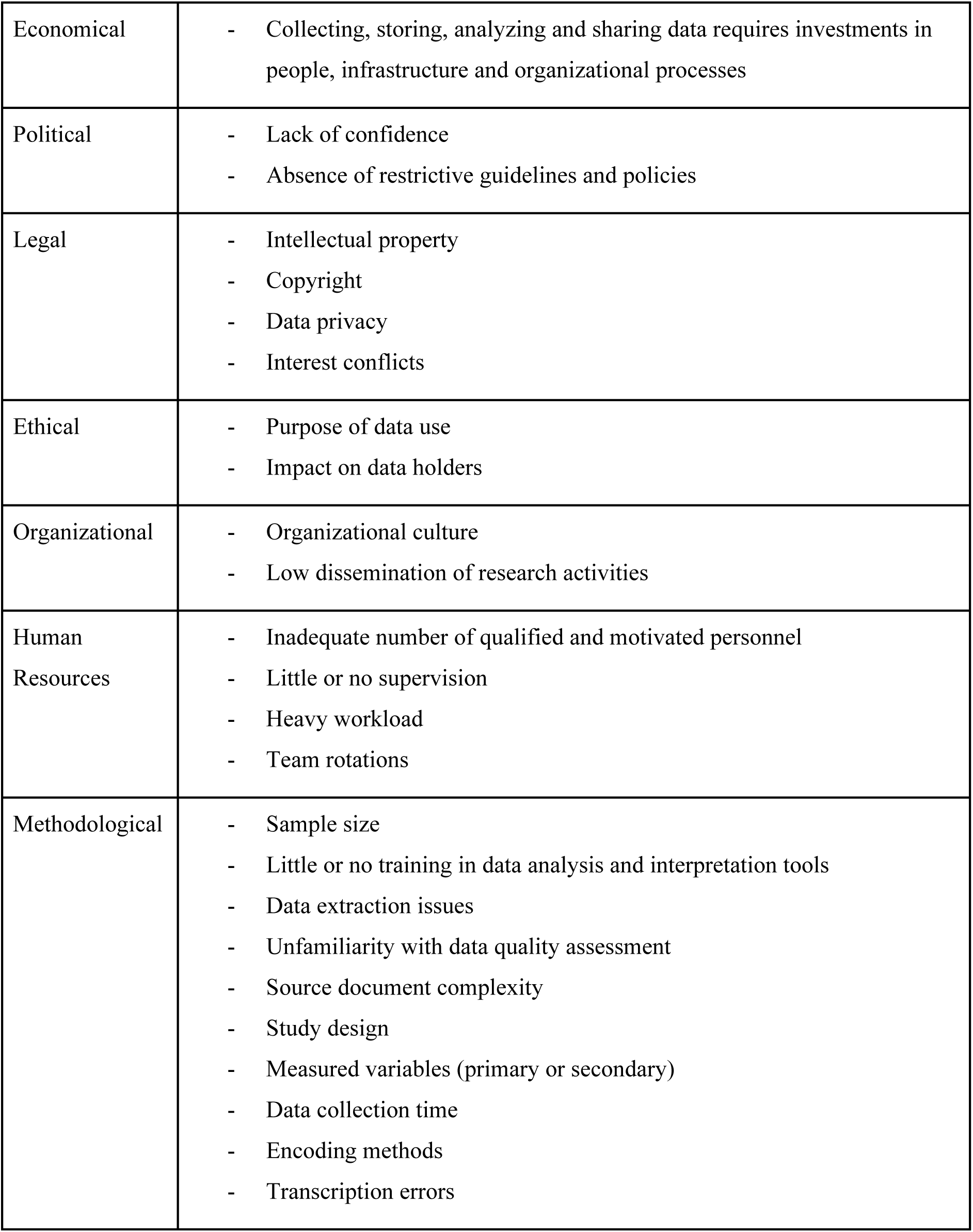
Barriers and factors for health data quality Source: Prepared by the author.

| Barrier | Factors |
| --- | --- |
| Technical | - Restrictive data formats<br>- Lack of metadata and standards<br>- Absence of technical solutions (e.g., interoperability)<br>- Poor design quality (standards) development (flexibility) and evaluation (usability and complexity) of system designs<br>- Lack of detailed information for specific searches<br>- Terminology variations<br>- Limited recovery capabilities<br>- Large amount of unstructured data;<br>- Challenges with patient identification and matching |
| Motivational | - Lack of incentives to use data in decision making<br>- Lack of delegation of responsibilities<br>- Lack of clarity of role and data owners |
| Economical | - Collecting, storing, analyzing and sharing data requires investments in people, infrastructure and organizational processes |
| Political | - Lack of confidence<br>- Absence of restrictive guidelines and policies |
| Legal | - Intellectual property<br>- Copyright<br>- Data privacy<br>- Interest conflicts |
| Ethical | - Purpose of data use<br>- Impact on data holders |
| Organizational | - Organizational culture<br>- Low dissemination of research activities |
| Human Resources | - Inadequate number of qualified and motivated personnel<br>- Little or no supervision<br>- Heavy workload<br>- Team rotations |
| Methodological | - Sample size<br>- Little or no training in data analysis and interpretation tools<br>- Data extraction issues<br>- Unfamiliarity with data quality assessment<br>- Source document complexity<br>- Study design<br>- Measured variables (primary or secondary)<br>- Data collection time<br>- Encoding methods<br>- Transcription errors |

Source: Prepared by the author.

Although many electronic records provide a dictionary of data from their sources, units of measurement are often neglected and adopted outside of established standards. Such “human errors” are inevitable, which reinforces the need for continuous quality assessment from the beginning of its collection. On the other hand, some studies have tried to develop ontologies in order to allow the automated and reproducible calculation of data quality measures. However, this strategy did not have great acceptance. For Feder [21], “the harmonized data quality assessment terminology, although not comprehensive, covers common and important aspects of the quality assessment practice”. Therefore, the generation of a data dictionary with its determined types and the creation of a data management plan are fundamental in the planning of research [22].

Both the way of collecting and inputting data impact the expected result of a dataset. Therefore, with a focus on minimizing data entry errors as an essential control strategy for clinical research studies, the implementation of intervention modes of technical barriers was presented, in the form of pre and post analysis [23]. The problems were caused by errors in the data source, extraction, transform, load process, or by limitations of the data entry tool. Extracting information to identify actionable insights by mining clinical documents can help answer quantitative questions derived from structured health quality research data sources [24].

Typical manual curation was considered insufficient given the time and effort involved in the iterative error detection process. The main sources of error included human and technological errors [25]. However, outliers identified by automated algorithms should be considered as potential outliers, leaving the field specialists in charge [26]. On the other hand, different and ambiguous definitions for the dimensions of data quality and related characteristics in the field of emergency medical services were presented [21]. Such divergences were based on intuition, previous experiences, and evaluation purposes. The use of definitions based on ontology or standardization is suggested for the purpose of comparing research methods and their results. The definitions and relationships between the different dimensions of data quality are not clear, which makes the quality of comparative assessment difficult [27].

In terms of evaluation methods, similar definitions overlap. The difference lies in the concept of distribution comparison and validity verification, where the definition of distribution comparison is based on the comparison of a data element with an official external reference [28]. Meanwhile, the validity check is concerned that a certain value is an outlier, that is, a value outside the normal range. The reasons for the existence of multiple evaluation practices are due to the heterogeneity of data sources in relation to syntax (file format), schema (data structure models), and semantics (meaning and varied interpretations) [17]. It is desirable that there is a common set of data to deal with such inconsistencies and allow the transformation of data into a structure capable of interoperating with its electronic records [29].

Data standardization transforms databases from disparate sources into a common format, with shared specifications and structures. It also allows users from different institutions to share digital resources and can facilitate the merging of multicenter data and the development of federated research networks [30]. For this, two processes are necessary: 1) the standardization of individual data elements, adhering to terminology specifications [31]; 2) standardizing the database structure through a CMD, which specifies where data values are located and stored in the database [32]. Improvements in electronic collection software functionality and its coding structures have also been reported to result in lower error rates [33]. In addition, it is recommended to have knowledge of the study platform and access to secondary data sources that can be used. In this way, transparency in the systemic dissemination of data quality with clear communication, well-defined processes, and instruments, can improve the multidisciplinary cooperation that the area requires [34]. Awareness campaigns on the topic at the organizational level also contributed to improving aspects of data governance. The most-reported error prevention activities were the continuing education of professionals with regular training of data collectors during their studies [32]. In this sense, in-service education should promote the correct use of names formulated by structured systems to improve the consistency and accuracy of records and favor their regular auditing. Health systems that received financial incentives for their research obtained more satisfactory results regarding the degree of reliability of their data [35].

## Discussion

The ultimate goal of data quality is to develop a disciplined process of identifying data sources, preparing the data for use, and evaluating the value of those sources for their intended use. In this sense, in addition to mapping concepts between different sources and application scenarios, it is essential to understand how initial data quality approaches are anchored in previous concepts and domains, with significant attention to suitability for use, following guidelines or use frameworks in a given context [20]. Thus, since the concept in the same data source can change over time, it is still necessary to carry out mapping with an emphasis on its dimensions in a judicious way and on how the evolution of concepts, processes and tools impact the quality assessment. of research and health services [36].

We know that the landscape of data quality research and its practical applications has changed rapidly and, therefore, the large amount and dissonance between domain definitions has increasingly motivated the search for a gold standard to be followed [37]. Widely considered anecdotal and esoteric, the area has received special attention, especially after the term “Big Data” gained more and more strength [38]. With the rapid growth of the field of data analysis in recent years, the quality of data has become increasingly significant and the field of research has witnessed extraordinary growth [3], mainly driven by digital solutions arising from the need for control. and the human inability to act with a large volume of information in research.

For example, recent clinical and health service research has adopted the concept of “fit for use” proposed in the information science literature. This concept implies that data quality dimensions do not have objective definitions, but are dependent on a set of tasks characterized by research methods and processes [39]. Thus, increasingly, data quality research has borrowed concepts from various referencing disciplines and, more importantly, with many different referencing disciplines using data quality as a context within their own discipline, the identity of the field. of research has become less and less distinct [40].

This transdisciplinarity may be reaching the threshold of significant growth and thus forcing the need for a metamorphosis of the area from focusing on the measurement and evaluation of data quality, today focused on content, to a direction focused on use and context [38]. In its broadest sense, the topic transcends all disciplinary boundaries of research activities. Therefore, creative approaches to decision-making in data quality and data usability require good use of transdisciplinary, regardless of study design planning and/or application area [41]. In the absence of a standard definition, the use of the concept “fit for purpose” for performance monitoring, program management, and data quality decision making has been increasingly used. As a large part of this quality is dependent on the collection stage, interventions must target the local level where it occurs, that is, they must encompass professionals at the operational level and forms at the technical level [22].

Thus, it is also critical to identify and address behavioral and organizational challenges, along with building technical capacity, increasingly fostering a “data-driven culture”. Yet, as a consequence of this lack of standard, the use of integrated quality assurance methods combined with standard operating procedures (SOPs), the use of rapid data feedback, and supportive supervision during the implementation of the surveys are feasible, effective, and necessary to ensure high-quality data [42,43].

Although data quality tools are defined from the end-user perspective, they are generally implemented from the researcher’s perspective [40]. A dataset is highly context-specific, i.e., a generic assessment framework is unlikely to provide a comprehensive analysis of data quality for a specific study, which makes its selection dependent on the analysis plan for the study [29].

The use of ontologies, for example, can help quantify the impact of likely problems and promote the validity of an effective electronic measure, allowing the assessment approach to be generalized to other data analysis tasks in more specific domains [23]. This benefit allows helping the decision-making process and planning of corrective actions and resources allocation faster [36].

On the other hand, the complex coding process can generate inconsistencies and incompleteness due to the characterization of clinically significant conditions, low clinical documentation and variability in interpretation [44]. Therefore, it is critical to use specific rules that capture relevant associations in their corresponding information groups. It is further suggested that administrative health data can capture valuable information about such difficulties using standardized terminologies and monitor and compare coded data between institutions [17].

The adoption of such well-defined interventions still plays an important role in data quality management. Such activity is performed through methods of process control and monitoring, tools that allow the manipulation and visualization of data in an effective way, in addition to carrying out experiments through techniques and analysis as a fundamental element in the discovery of patterns and perspectives on the target information subset [45].

Even when such information is available, there are still some aspects that hinder its use by researchers, managers and health professionals, such as the non-computerization of processes, the heterogeneity of isolated data silos, duplicity and errors in data collection and processing. in health information systems. Consequently, there is a decrease in the quality of information, which makes its coordination, evaluation and reproduction difficult [46].

Despite the intense volume of production, information still remains decentralized without being able to help the decision-making process of planning many types of research [47]. In this way, the reuse of information, due to the lack of reliability and standardization of quality mechanisms in subsequent research on a given topic, has a great negative impact [39]. On the other hand, the centralized storage of variables without an adequate mapping to changes in system paradigms (metadata) and with a mechanism to trace the effects of changes in concepts that are frequent in the health area, can also affect the degree of reliability of research [48]. For example, the definition of severity classification of a given condition can change over time, and consequently, mitigate the comparability power of a study, or even prevent it from being used as a basis for planning or evaluating a new one [28].

Therefore, as well as the concerns of the pillars of philosophy (of being, knowledge, and language), the realization of mapping with emphasis on domains or concepts must coexist in health information systems so that your faculty has the intention to favor the maximization of processes, increase their productivity, reduce costs and meet research needs [49]. Consequently, within legal and ethical limits, it is increasingly necessary to use data in the most comprehensive and efficient way possible for the benefit of patients [50].

Regardless of the model adopted, these tools should aim to discover abnormalities and the ability to stop and correct them in an acceptable time, also allowing for the investigation of the cause of the problem [23]. This allows the results and processes to contribute to the quality and, therefore, to a more efficient search. Technology is a great ally in production processes, and in parallel with the tools of the Lean Six Sigma philosophy, they can partially replace human work [51].

To maximize the potential of this pillar, the value derived from using analytics must dictate data quality requirements. In this sense, fields such as artificial intelligence have been increasingly adopted. The use of Computer Vision/Deep Learning, for example, a technology to visualize multidimensional data, has demonstrated data quality checks with a systematic approach, in order to guarantee a reliable and viable developed asset for healthcare organizations for the holistic implementation of machine learning processes [35]. However, most of these analytical tools still assume that the analyzed data have high intrinsic quality, which can thus allow possible failures in the process, in addition to the lack of optimization, safety and reliability of the final experiments [48].

Important research in the area of RD has been carried out in this regard, with regard not only to the quality of the content of a record but also to the quality of how the data and what mechanisms are used to make them available for wider use [20]. As outlined in Recommendations for Improving the Quality of Rare Disease Registries, managing data at the source and applying the FAIR Guiding Principles for its management are recognized as fundamental strategies in collaboration. interdisciplinary research network [52].

It is known that the cultural background and experience of researchers can influence the interpretation of data [34]. Therefore, for decentralized research networks, a combination of integrated tools located both centrally and at each partner site can increase the quality of research data [29]. A central metadata repository contains common data elements and value definitions that are used to validate the content of data warehouses operated at each location [30]. In this way, the consortium can work with standardized reports on data quality, preserving the autonomy of each partner site and allowing individual centers to improve data in their locally sourced systems [51].

In addition, the shared use and promotion of training of health professionals in data management can facilitate the conduct of more quality studies in low- and middle-income countries. In turn, the low level of use of health information and poor management of health information systems presented in these countries make evidence-based decisions and planning at the community level difficult [2].

Although poor practices and investment needs in health information systems have been found, improved delivery of health services with greater efficiency based on health data has been reported. The results also demonstrate that, despite existing, such individual training efforts focus mainly on the transmission of data analysis skills [40]. This implies that the effect on the performance of health systems and research in these countries still has major challenges at strategic stages such as planning and managing complete data, leading to errors in population health management and clinical care [19].

The production of data and dissemination of quality information, therefore, depend on establishing a record governance model, identifying the correct data sources, specifying data elements, case report forms, standardization and building an information technology infrastructure in accordance with agreed principles [51]. Other topics, such as developing adequate documentation, training staff and providing audit data quality, are also considered essential to improve the quality of the record and serve as a reference for teaching material for health service education [47].

Once established, the governance model combined with the operationalization of elements that incorporate activities of periodic verification, such as the duplicity of participants and remote monitoring, it is therefore expected that there will be the greater speed of processes and minimization of risks to the integrity of the studies [25]. A good governance model must also mutually respect the ethical precepts and the internal and procedural policies of the institutions that make up the research [19].

The paradox of how to guarantee the unique access of data and consequently its reliability for consumption in analysis and artificial intelligence algorithms must still walk respecting the laws and good practices of protection of sensitive data

There are significant differences between the development of health and AI, which suggest to the latter that a principled approach is different from that already adopted by health [36]. In parallel, the challenges of AI concern the establishment of shared purposes, assurance mechanisms, specific history and guidelines, proven methods for translating principles into practice and robust mechanisms of legal and professional accountability [50].

Clearly, there is a need to take a proactive ‘digital ethics’ approach to these challenges, so that the transition from an assessment of what is morally good to what is politically feasible and legally feasible takes place before ethical mistakes lead to social rejection [53], and leave institutions and researchers unable to benefit from the so-called ‘double advantage’, where opportunities are capitalized and risks mitigated [50] of an ethical approach to governance [53].

## Conclusion

This study should help researchers, data managers, auditors and systems engineers to think about the conception, monitoring, tools and methodologies used to design, execute and evaluate their researches and proposals that are concerned with data quality. With a well-established and validated data quality workflow for healthcare, we hope to also assist in mapping the management processes of healthcare research and promote the identification of gaps in the collection flow where any necessary data quality intervention can be evaluated accordingly with the best tools described here.

Identifying systematic and persistent defects in advance and correctly directing human, technical and financial resources are essential to promote better management and increase the quality of information and results achieved in research. Indirectly, this step can provide improvements and benefits to health managers, allowing greater efficiency in services, better allocation of resources and promoting benefits to society through relevant data that impact the performance and effectiveness of public health services, as well as boost areas of research, innovation and enterprise development.

Among the limitations of our review, we highlight the search for works written in English and Portuguese, since the interpretation of concepts and even the translations of terms referring to the dimensions and adaptations to different cultural realities can vary and thus influence part of our evaluation [43]. In addition, the absence of evidence in emerging countries did not allow us to carry out an adequate synthesis regarding the performance and application of the evidence found [2]. Finally, due to the rapid growth of technologies applied to the area, we carried out a search focused on the last five years, which may abstract other fundamentals and relevant procedures.

The routine of health services that deal with demands for collecting and consuming data and information can benefit from the set of evidence on tools, processes and evaluation techniques presented here. Increasingly ubiquitous in the daily lives of professionals, managers and patients, technology should not be adopted in an anecdotal way, as such conduct can generate misinterpreted information obtained from unreliable digital health devices and systems.

In this way, the resources presented can help guide medical decisions that do not only involve medical professionals, and indirectly contribute to avoiding decisions based on low-quality information that can put patients’ lives at risk. Thus, with the effect of elucidating health institutions that focus on a scenario that promotes a data culture and, consequently, are qualified to provide quality information, it is expected that the mechanisms presented can also enable better conditions for patient care.

Once the technical and organizational barriers have been overcome, with data managed, reused, stored, extracted and distributed properly [53], health must also pay attention to behavior focused on interactions between human, artificial and hybrid actors. This reflects the importance of adhering to social, ethical and professional norms in the inclusion of demands related to justice, responsibility and transparency [54]. In short, increasing dependence on “quality information” increases its possibilities [53], but it also presents regulators and policymakers with considerable challenges related to their governance in health.

With the promotion of the data culture increasingly present in a transversal way, we believe that research and researchers can offer more and more reliable evidence and in this way benefit the promotion and approach to the health area. This mutual cycle must be transparent so that the awareness that its adherence to such a practice can favor the potential strengthening of a collaborative network based on results, promote fluidity and methodological transparency, in addition to promoting the incentive to share and consequently the reuse of data into reliable information silos, thus enhancing the development and credibility of health research.

At the international level, the platforms that have a centralized structure of reliable data repositories in patients records that offer data sharing have reduced duplication of efforts and costs. This collaboration can further decrease disparate inequities between emerging and developed countries, in addition to giving celerity and minimizing risks in the development and integrity of studies.

It is suggested as future work the development of a toolkit based on process indicators to verify the quality of existing records and provide a score and feedback on the aspects of the registry that require improvements. In conclusion, in the field of records, there is a need for coordination between undergoing initiatives at national and international levels. At the national level, we recommend the development of a centralized, public, national, “registration as a service” platform, which will guarantee access to highly trained personnel on all topics mentioned in the article, promoting the standardization of registries. In addition to allowing cost and time savings in creating new registries, the strategy should allow for the linking of important data sources on different diseases and increase the capacity to develop cooperation at the regional level.

In conclusion, we expect the results to provide evidence of best practices using data quality approaches, which involve many other stakeholders, not just researchers and research networks. Also, the information collected in this study can support better decision-making in the area and provide subsidies for the planning of health information policies. For future work, we can mention the use of the data models found in this study to serve as a structured information base for decision-support information systems development and health observatories, which are increasingly relevant to public health. Furthermore, concerning the health context, it may allow the execution of implementation research projects and the combination with frameworks that relate to health behavior interventions, for example, the RE-AIM framework [55], among others.

## Data Availability

The list of all findings included in our review is available at Supplementary file 1 - Individual studies and https://docs.google.com/spreadsheets/d/1l-1do1xn1jGq4uXrfHQrdnA_Y2LC_ku5/edit?usp=sharing&ouid=110857452492520611792&rtpof=true&sd=true:

